# Effect of Change Potassium Intake on Systolic Blood Pressure: A Dose-Response Meta-Analysis of Randomized Clinical Trials (2000-2024)

**DOI:** 10.1101/2024.11.27.24318119

**Authors:** Maelys Granal, Victoria Sourd, Michel Burnier, Jean Pierre Fauvel, Arthur Gougeon

**Author notes:** **Corresponding author:** Maelys GRANAL, Evaluation et modélisation des effets thérapeutiques – Laboratoire de biométrie et biologie évolutive – UMR 5558, Université Lyon 1 site Laennec, Hospices Civils de Lyon, Hôpital Edouard Herriot, Service de Néphrologie.

## Abstract

**Background:** The global prevalence of hypertension in adults has doubled in 30 years, rising from 650 million to 1.3 billion between 1990 and 2019. Although reducing sodium intake is one of the first core of non-pharmacological hypertension management guidelines, recent guidelines strongly emphasize the need for increased potassium intake due to accumulating evidence of its cardiovascular benefits.

**Methods:** We conducted a systematic review to identify randomized controlled trials evaluating the effect of potassium supplementation (measured through 24-hour urinary potassium excretion) on systolic blood pressure. A dose-response meta-analysis was performed using three regression models: linear, quadratic and one-stage cubic spline. Subgroup analyses were performed according to hypertensive or normotensive status.

**Results:** The meta-analysis included 10 randomized clinical trials comprising 4 studies in normotensive individuals and 6 in hypertensive patients. The dose-response relationship differed based on participants’ blood pressure status. Subgroup analyses revealed a weak negative linear effect of potassium on systolic blood pressure in normotensive individuals, while a stronger negative linear relationship was observed in hypertensive patients. The dose-response meta-analysis predicted that for a 50 mmol increase in urinary potassium excretion over 24 hours, systolic blood pressure decreased by 0.5 mmHg in the normotensive group and by 5.3 mmHg in the hypertensive group.

**Conclusion:** These meta-analysis results confirm a dose-response relationship between potassium supplementation and systolic blood pressure reduction, particularly in individuals with hypertension. Predictions should be viewed with caution due to the limited number of studies included in the study.

**Novelty and relevance:** *What is new?:*

- The dose-response relationship between change in potassium intake and change in systolic blood pressure was described using three statistical models (linear, quadratic and cubic spline) to avoid *a priori* assumptions.
- The dose-response meta-analysis revealed a weak linear relationship between increased potassium intake and decreased systolic blood pressure in normotensive individuals. The effect was more pronounced in hypertensive patients.

*What is relevant?:*

- The meta-analysis specifically addresses the impact of change in potassium intake on change in systolic blood pressure selecting only recent (since the 2000s), high-quality randomized clinical trials in witch potassium intake was accurately estimated through 24-hour urinary potassium excretion. The included studies underwent rigorous assessment of bias. These analyses stratified by normotensive or hypertensive status align with current therapeutic and nutritional management practices, as well as recent diagnostic methods (measuring devices, thresholds). These findings aim to facilitate decision-making for future national and international nutritional recommendations.

*Clinical/pathophysiological implications?:*

- The results of this dose-response meta-analysis support current nutritional recommendations to increase dietary potassium intake to lower blood pressure, particularly in hypertensive patients. Improved potassium intake management could significantly improve blood pressure control and reduce cardiovascular risk, particularly in high-risk hypertensive patients.

## Introduction

Cardiovascular diseases (CVD) remain the leading cause of mortality and disability worldwide, with an estimated 18.6 million deaths and 393 million years of disability each year^1^. Hypertension is the primary risk factor for CVD^2–4^. It is estimated that reducing the population’s blood pressure (BP) distribution by 5 mmHg could prevent one-third of strokes and one-fifth of coronary events^5^. Non-pharmacological interventions, such as dietary and lifestyle modifications, are fundamental to hypertension management. Indeed, a close link has been established between nutrition and BP, with the identification of various factors likely to reduce BP, including weight reduction, moderation of alcohol consumption, reduction of sodium intake and increase in potassium intake^6–9^. These non-pharmacological measures to lower BP are widely recognized in all national and international dietary recommendations and guidelines^10–16^ although they can be challenging to implement in clinical practice. Despite reducing sodium intake has long been a cornerstone of hypertension management, recent guidelines have underline the need to increase potassium intake based on recent trials^17,18^. The World Health Organization (WHO)^14^, the AHA^19^ and the International and European hypertension guidelines^13,15,16^ recommend a dietary potassium intake exceeding 3.5 g/day (∼90 mmol/day). The benefits of a potassium-enriched diet in reducing cardiovascular risk have been the topic of numerous publications and it has even been established that a higher intake of potassium was associated with a reduction of BP and a lower risk of stroke^19–30^. Non-pharmacological interventions to reduce BP are therefore a top priority for both patients and healthcare providers^31^. They not only improve quality of life by reducing the need for therapeutic management^32^, they also promote overall health through balanced dietary habits. The beneficial effects of nutrition on cardiovascular risk also enable the patient to gain in quantity of life^33,34^. Additionally, these measures have the potential to reduce healthcare costs associated with antihypertensive medications, hospitalizations, and management of CVD-related complications. Previous dose-response meta-analysis by Filippini et al. (2017^35^ and 2020^36^) examined the relationship between potassium intake and SBP reporting a significant correlation between the two parameters^36^. Like many other earlier meta-analyses^37–39^, their review included studies conducted prior 2000, which may not fully reflect current BP measurements, therapeutic management practices or dietary recommendations^15,40–42^. Given the significant evolution of hypertension guidelines over the past two decades - particularly of the increased emphasis on potassium intake - we hypothesized that a meta-analysis based on more recent data and limited to randomized clinical trials (RCT) devoted to the effects of systolic BP (SBP) related to changes in potassium intake alone (since modifications in sodium and potassium intake have interrelated effects) would provide more recent and relevant insights, in line with current practice.

The aim of this study was to perform a dose-response meta-analysis of changes in potassium intake alone (assessed by 24-hour urinary potassium excretion) with changes in SBP, using recent high-quality RCTs, focusing exclusively on post-2000 data. Consistent with current clinical and dietary practices, this approach aims to refine and validate potassium intake recommendations for hypertension management.

## Methods

### Protocol and registration

The protocol was registered *a priori* on Prospero: CRD42023440909. The methodology complied with the Cochrane Handbook for Systematic Reviews of interventions ^43^ and the study was reported in accordance with the PRISMA checklist^44^ (see, supplementary material S1).

### Search strategy and study selection

A systematic literature search was conducted in PubMed, Cochrane Central and Embase up to September 2024. Our PICOT criteria were as follows: (P) humans, (I) modification of potassium intake, (C) no or other modification of potassium intake, (O) Systolic Blood Pressure, (T) systematic reviews and meta-analysis. The search strategy was a combination of different keywords for each criterion (for details see, supplementary material S2]). The search was restricted to articles published in English and French. The research focused on systematic reviews and meta-analyses, as numerous meta-analyses had already been conducted on this topic. Additionally, a manual search of references was carried out.

Eligibility was assessed independently by two independent reviewers (MG and VS). The records were screen by title and abstract, and subsequently in Full-text based on PICOT (MG and VS). Any disagreements were resolved by a third reviewer (AG). Only studies estimating potassium intake via 24-hour urinary excretion and published after January 1, 2000, were included to ensure methodological consistency with contemporary BP measurement and management practices of therapeutic and dietary. Studies involving sodium-only interventions, sodium and potassium associated interventions (salt substitute, diet intervention) or pediatric populations were excluded. Authors of studies with incomplete data were contacted, and studies were excluded if no response was received.

To avoid overlap in data extraction, a correspondence table was created to compare the original articles included in the meta-analyses. Clinical studies included in several systematic reviews were considered only once.

### Data extraction

Two independent reviewers (MG, VS) extracted the data using a standardized form. When data of a clinical study was missing in the included systematic reviews, we extracted them from the RCT study. Extracted data included: study characteristics (study ID, author, date of publication, country, study design, washout period, number of participants in the intervention and control groups, duration of intervention), population characteristics (age, proportion of female participants, pathology population, CKD population, estimated glomerular filtration rate, hypertension status, body mass index (BMI)), outcome data (BP measurement method, baseline and follow up SBP, and measured 24-hour urinary potassium excretion).

For studies not reporting the Standard Deviation (SD) of means for each group, these were converted from standard errors (SE), median interquartile range, or 95% confidence interval (CI95). For studies that used a crossover design, the effect of the intervention was taken as the difference between endpoint measurements in the two groups. For studies that used a parallel design, we did not adjust for baseline differences in SBP. It was assumed that randomization would balance baseline characteristics between groups, and any differences would be minimal and unlikely to significantly affect the measured intervention effects. The duration of the intervention was defined as the period from the start of the intervention to the point at which the outcome was measured. Ambulatory SBP measurements were prioritized over clinic readings, and daytime SBP was prioritized over nighttime SBP.

### Statistical Analysis

A random-effects dose-response meta-analysis was performed, with potassium dose expressed as the standardized mean difference (SMD) in urinary potassium excretion between low and high-intake groups., with the low potassium group referenced at 0. The response was defined as the mean difference in SBP between the groups at the end of the intervention.

Three regression models (linear, quadratic, and cubic spline) were used to assess the dose-response relationship. The cubic spline model included knots at the 15th, 50th, and 85^th^ percentiles of potassium intake distribution. Model selection was based on the Akaike Information Criterion (AIC), and the lowest AIC was considered the best fit for the data. A table of AIC values and dose-response curves for the different models by population is available (see, supplementary material S3). The selected model was used to predict the dose-response relationship for potassium intake values ranging from 10 mmol/day to 100 mmol/day.

Given the limited number of studies included in the present meta-analysis, sensitivity analyses were not performed based on study design or intervention duration, as originally planned. Only a subgroup analysis in hypertensive or normotensive subjects was performed.

Publication bias was assessed using funnel plot and Egger’s test^45^, independently reviewed by two independent reviewers (MG and AG) Analyses were conducted using the R 4.2.2 software^46^ and the *dosresmeta* and *metafor*^47^ *packages*.

All p-values were considered significant at 0.05 without multiple testing adjustment.

### Risk of bias assessment

The Cochrane ’’Risk of Bias’’ 2.0 tool ^48,49^ was employed to evaluate the risk of bias of the included studies by MG across 5 domains: randomization process, deviation from planned intervention, missing outcome data, outcome measurement and outcome reporting. Each domain was classified as “low risk of bias”, “concern” or “high risk” based on responses to the guideline questions.

## Results

### Study selection and characteristics

Out of the 1,826 identified references, 3 systematic reviews were included, encompassing a total of 10 RCT (Figure 1). These RCTs were published between 2005 and 2021. The majority of the studies focused on hypertensive patients (60%).

**Figure 1.**
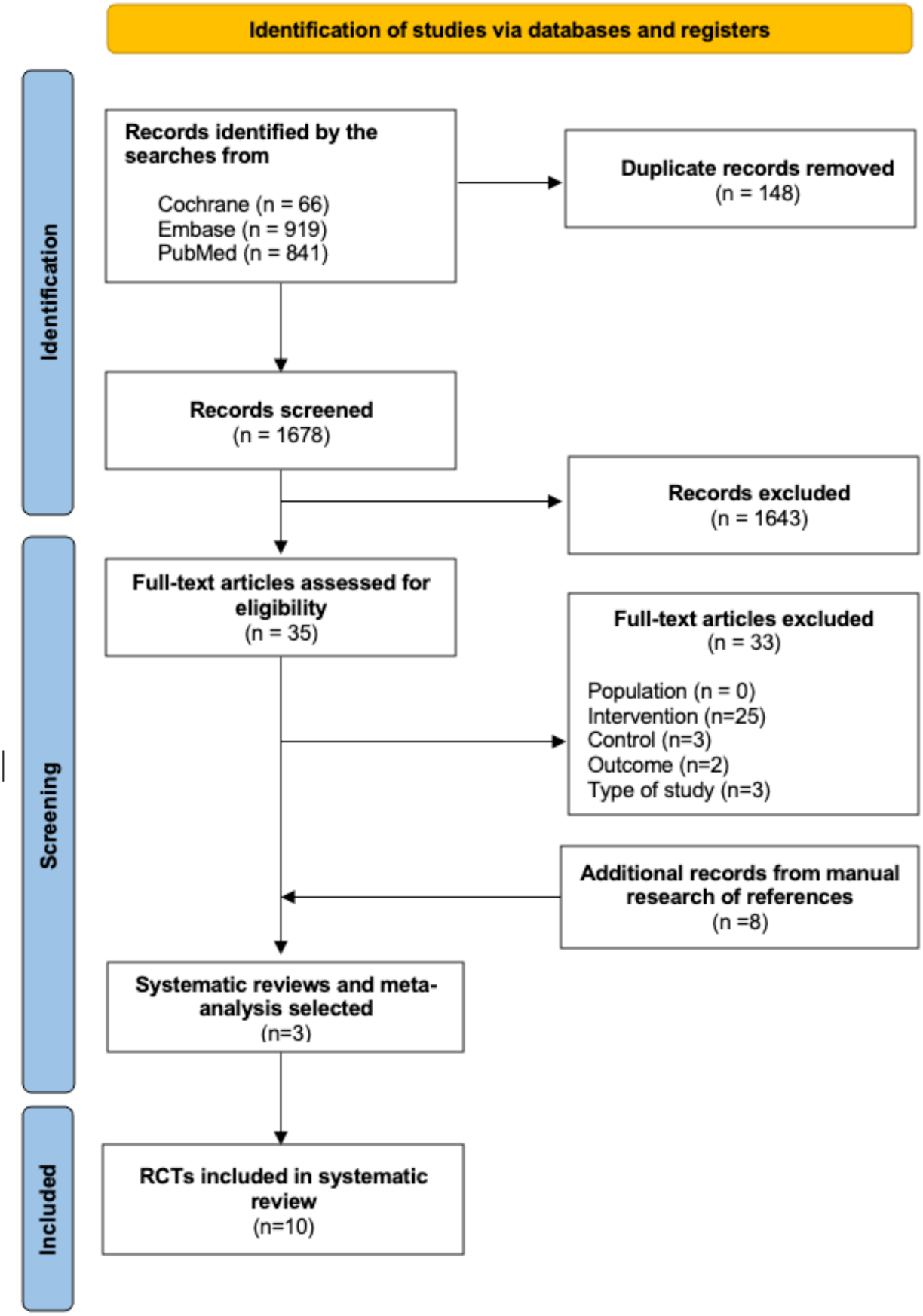
Flow diagram PRISMA of included RCTs in the meta-analysis

Table 1 shows the main characteristics of the 10 RCTs included in the dose-response meta-analysis. The studies were published between 2005 and 2021, involving a total of 684 patients, 342 in the intervention group (potassium intake modification) and 342 in the control group (unchanged diet). RCTs were conducted in adults in the UK (N=4), USA (N=1), Italy (N=1), China (N=1), Netherlands (N=1) and Denmark (N=2). Among the 10 RCTs, one was conducted in parallel, the others in crossover. Intervention durations ranged from 1 to 6 weeks. Blood pressure (BP) measurements were conducted in clinical settings in two studies, whereas the other eight employed ambulatory monitoring. Participant characteristics varied, with mean ages ranging from 26 to 66 years and mean BMIs between 22 and 31 kg/m². Most study groups were mixed, although two studies focused exclusively on male participants. Among the study groups, 4 study groups consisted of normotensive individuals, while 6 involved hypertensive patients. Subgroup analyses were conducted to assess outcomes based on participants’ normotensive or hypertensive status.

**Table 1.**
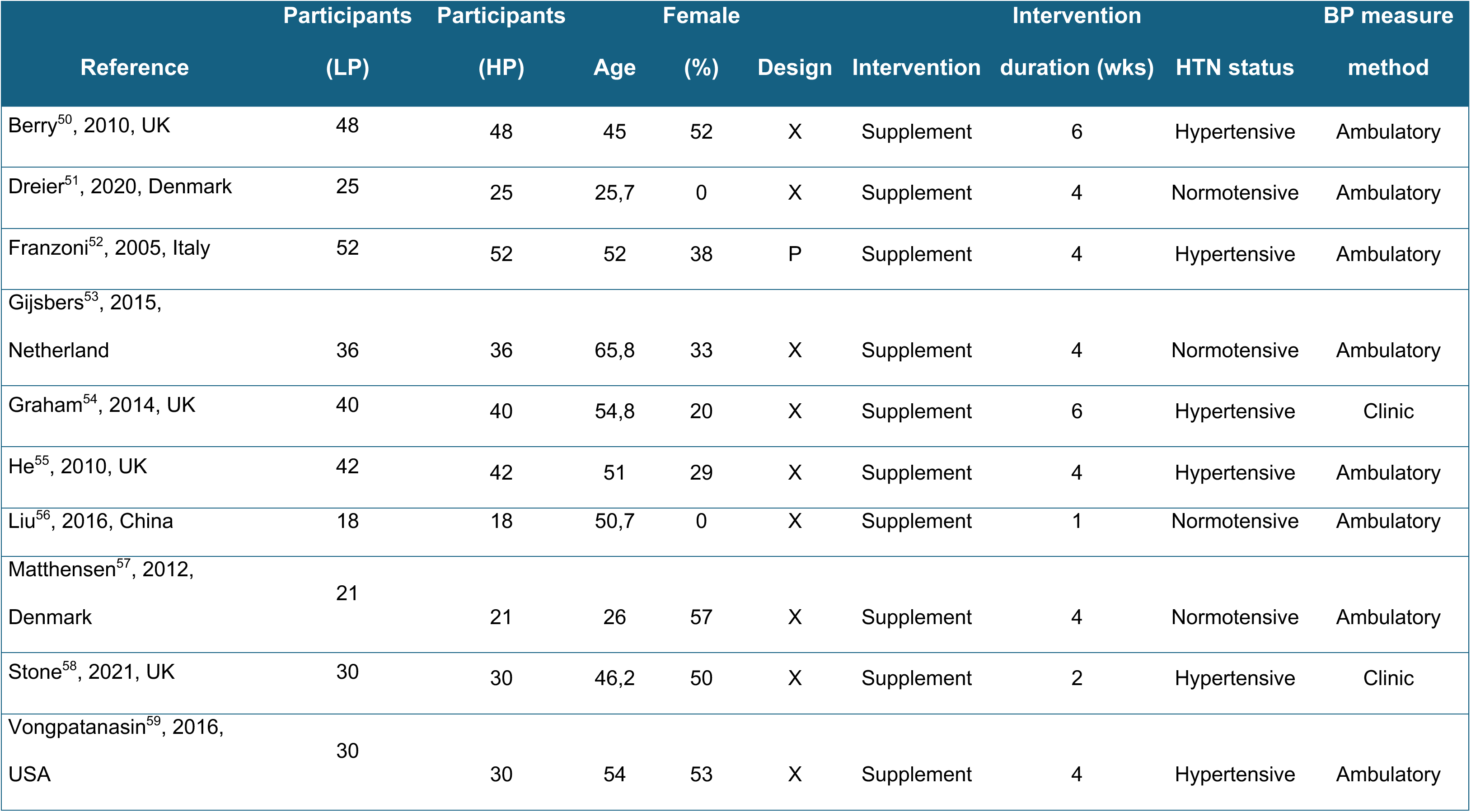
Characteristics of the 10 randomized clinical trials included.

### Description of potassium intake modification

The interventions were designed to increase potassium intake through supplementation with potassium chloride, potassium aspartate, potassium bicarbonate or increase consumption of potassium-rich fruits and vegetables. Dietary interventions occasionally included specific dietary instructions, patients were generally asked to maintain their usual dietary habits, except for the potassium supplementation, and to maintain their usual physical activities. In the high-potassium groups, supplements were administered as potassium tablets or slow-released potassium tablets. For crossover trials, participants were randomly assigned to groups, with some trials including a washout period between intervention phases. A run-in period was frequently used in both groups before administering potassium supplements versus placebo, to assess participants’ ability to adhere to the different dietary interventions.

In all included RCTs, 24-hour urinary potassium excretion and SBP were evaluated by a third party. BP measurements were performed at the beginning and end of the intervention for parallel studies, and at the end of each intervention phase in crossover studies.

### Dose-response meta-analysis

Standardized Mean differences (SMD) in potassium excretion ranged from 12.5 to 92 mmol/day across the overall population, with ranges of 30 to 92 mmol/day in normotensive patients and 12.5 to 45 mmol/day in hypertensive patients. In normotensives, a slight negative linear relationship was observed between increased potassium intake and changes in SBP. In contrast, a more pronounced negative linear relationship was evident in hypertensive patients (figure 2b and 2c). For instance, an increase of 24-hour urinary potassium excretion of 50 mmol/day the hypotensive effects were estimated at -5.3 mmHg in SBP in hypertensives compared to a hypotensive effect of only 0.5 mmHg in normotensives. For all populations combined, the dose-response curve appeared to be non-linear (cubic spline). A U-shaped relationship was observed between changes in 24-hour urinary potassium excretion and SBP (figure 2a) highlighting differential effects depending on the magnitude of potassium intake.

**Figure 2.**
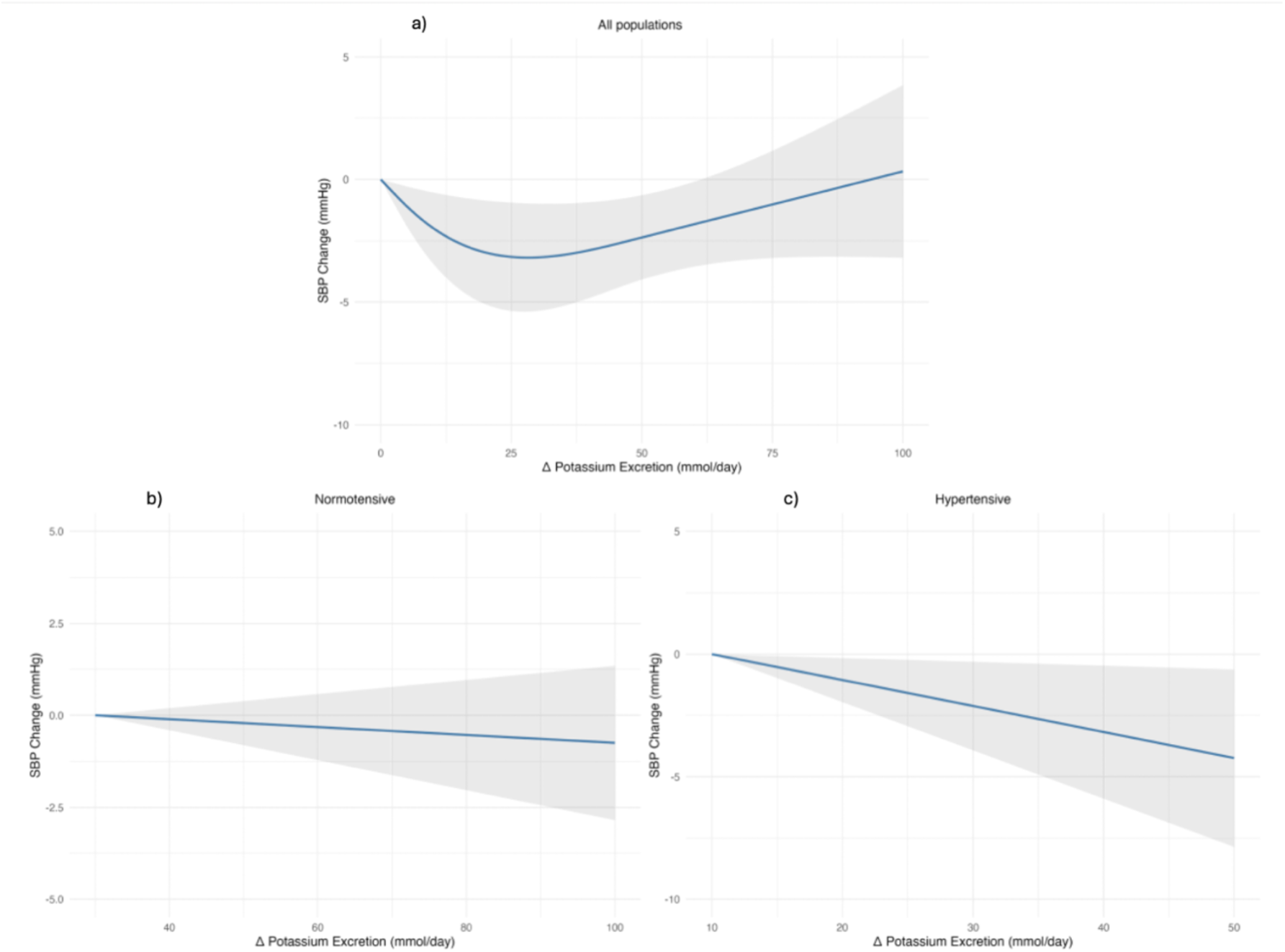
Dose-response meta-analysis of changes in systolic blood pressure (in mmHg) as a function of differences in urinary potassium excretion (in mmol/day) between the treatment group (potassium-supplemented group) and the control group with 95% confidence limits a) in all population (n=10), b) in normotensive population (n=4), c) in hypertensive population (n=6).

### Dose-response prediction

Predictions of changes in mean SBP as a function of changes in potassium intake of 10, 20, 30, 40, 50, 60, 70, 80, 90 and 100 mmol/day are summarized in Table 2. To minimize data extrapolation, SBP changes are presented within the range of 30 to 100 mmol/day change in potassium intake in normotensives and in the range of 10 to 50 mmol/day in hypertensives.

**Table 2.** Dose-response prediction of mean changes in systolic blood pressure (mmHg (CI95)) as a function of potassium dose (mmol/day). Linear models for the normotensive and the hypertensive dose-effect prediction. Cubic spline model for the all-groups dose-effect prediction

| Change in K intake<br>mmol/day | Change in SBP (mmHg) | Change in SBP (mmHg) | Change in SBP (mmHg) |
| --- | --- | --- | --- |
|  | Normotensives<br>(n=4 studies) | Hypertensives<br>(n=6 studies) | All studies‡<br>(n=10 studies) |
| 10 |  | -1.06 (-1.96 ; -0.16) | -1.98 (-3.43 ; -0.53) |
| 20 |  | -2.12 (-3.93 ; -0.31) | -2.97 (-5.09 ; -0.85) |
| 30 | -0.32 (-1.22 ; 0.58) | -3.18 (-5.89 ; -0.47) | -3.18 (-5.37 ; -0.99) |
| 40 | -0.43 (-1.63 ; 0.77) | -4.24 (-7.86 ; -0.63) | -2.88 (-4.83 ; -0.93) |
| 50 | -0.54 (-2.03 ; 0.96) | -5.30 (-9.82 ; -0.78) | -2.36 (-4.08 ; -0.64) |
| 60 | -0.64 (-2.44 ; 1.15) |  | -1.82 (-3.55 ; -0.10) |
| 70 | -0.75 (-2.84 ; 1.35) |  | -1.29 (-3.27 ; 0.70) |
| 80 | -0.85 (-3.25 ; 1.54) |  | -0.75 (-3.16 ; 1.67) |
| 90 | -0.96 (-3.66 ; 1.73) |  | -0.21 (-3.15 ; 2.73) |
| 100 | -1.07 (-4.06 ; 1.92) |  | 0.33 (-3.19 ; 3.85) |

### Risk of bias assessment

The risk-of-bias analysis, conducted across five domains: randomization process, deviation from the planned intervention, missing outcome data, outcome measurement, and outcome reporting, classified three studies as having a low overall risk of bias, while seven studies were categorized as raising concerns regarding the overall risk of bias. Four studies raised concerns about the blinding of participants or administrators. All studies were considered as low risk in terms of deviations from the planned interventions and missing outcome data. In contrast, one study was deemed to be of concern in terms of “outcome measurement”, as outcome assessors were likely to have been aware of the intervention received by study participants. Despite this, the likelihood of this knowledge influencing the results was deemed minimal. Half of the included studies raised concerns regarding the “selection of the reported outcome”, as it was not specified whether the data that produced this outcome were eligible for multiple analyses (figure 3). Detailed information regarding the risk of bias for each included study is provided in the supplementary data.

**Figure 3.**
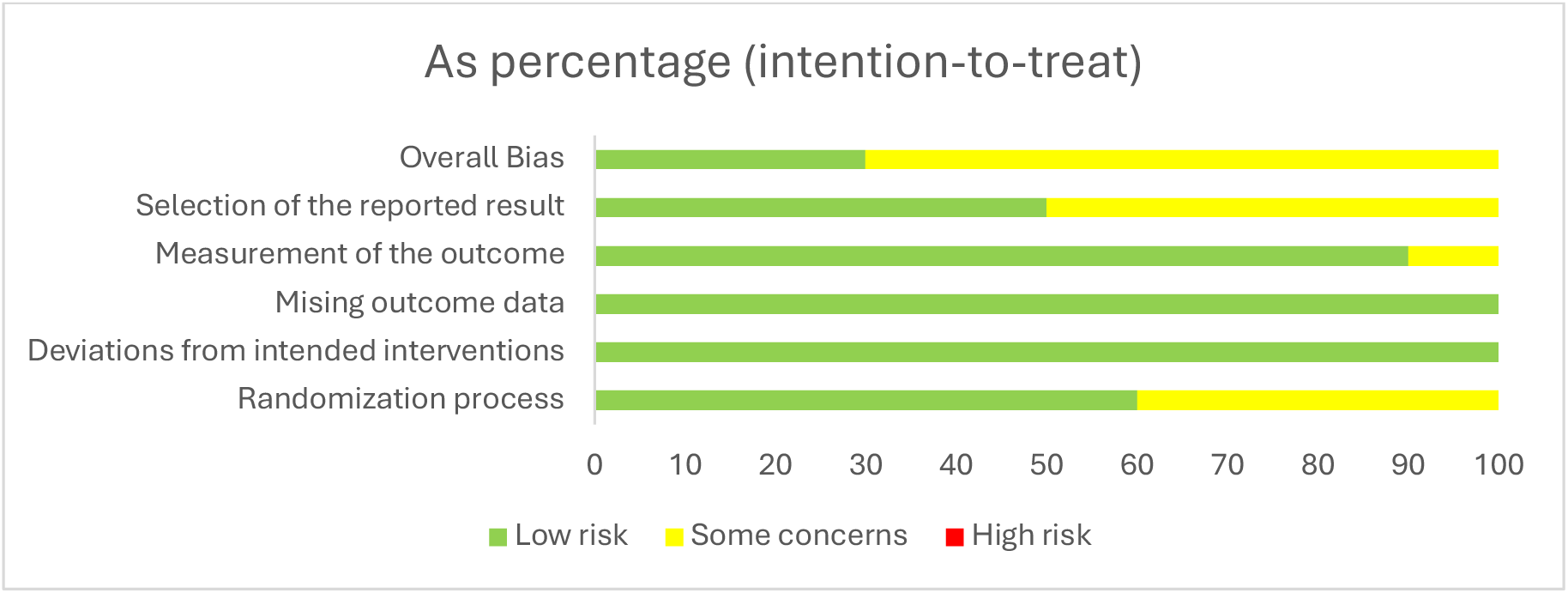
Summary of the risk of bias assessment

### Publication bias assessment

A visual inspection of the funnel plot did not indicate the presence of publication bias (Figure 4) This finding was further supported by the Egger’s test, which yielded a p=0.44 suggesting no significant publication bias

**Figure 4.**
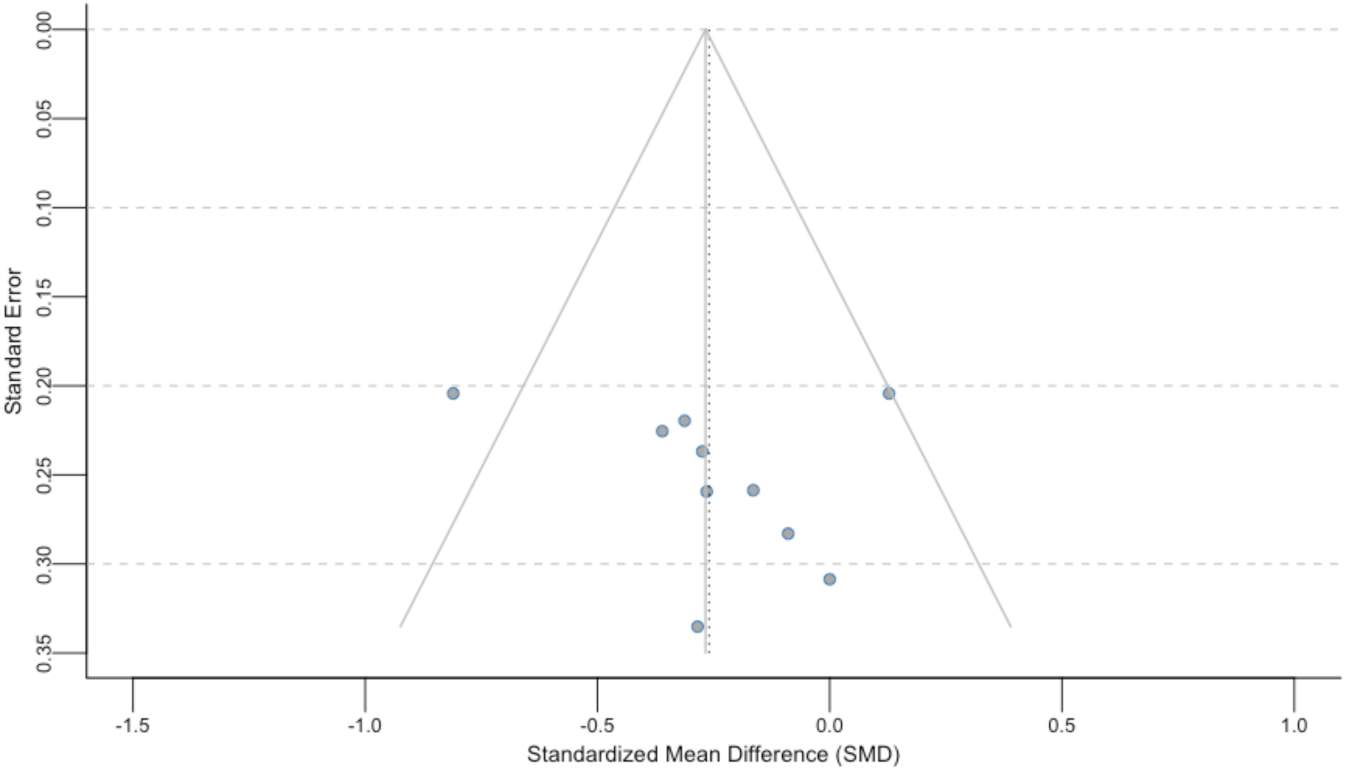
Funnel plot for publication bias for Standardized Mean Difference (SMD) for changes Systolic Blood Pressure (SBP) levels (as mmHg) and its standard error (SE).

## Discussion

We conducted a dose-response meta-analysis to evaluate the effect of changes in potassium intake on SBP focusing exclusively on studies published post-2000. This decision ensures consistency with contemporary dietary and therapeutic management of hypertension. Furthermore, the analysis was restricted to RCTs that reported 24- hour urinary potassium excretion to assess changes in potassium intake. This approach provides updated information to validate or refines current recommendations for potassium intake in the management of hypertension. Subgroup analyses support the negative relationship between increased potassium intake and decreased SBP in normotensives. This relationship was more pronounced in hypertensive individuals, aligning with findings from the previously published meta-analysis by *Filippini et al*^36^. The dose-response relationship between change in potassium intake and change in systolic blood pressure was best described using linear models in both groups, likely due to the limited number of studies included (normotensives: n = 4, hypertensives: n = 6), which restricted the ability to capture potential non-linear variations in effect. The U-shaped curve was observed in the overall population using a cubic spline model; however, this pattern is likely artefactual. Indeed, interventional studies reporting small increases in potassium intake (less than 50 mmol/day) with significant decreases in SBP were mainly carried out in hypertensive patients, and conversely, studies with a large variation in potassium intake (over 50 mmol/day) and which found a small decrease in SBP were carried out only in normotensives. The subgroup analysis aimed to differentiate the effects of potassium intake based on the normotensive or hypertensive status of participants, thus addressing the relevance of increasing potassium intake across populations or targeting those at higher cardiovascular risk, such as hypertensive patients. The results confirm that benefits of an increase in potassium intake using potassium tablets will mainly benefit to hypertensives^35,36^. In addition, hypertensive patients are likely to benefit more from additional potassium intake than normotensives, as they have a higher risk of CVD. Furthermore, dietary modifications to increase potassium intake (increased consumption of fruits and vegetables) offer additional benefits with fiber and vitamin supplementation for both normotensives and hypertensives^27^.

While the impact of increased potassium intake on BP reduction has already been the focus of several published meta-analyses^35,36,38,39,60^, the dose-response relationship has rarely been studied^36^. The strength of our study is that it provides a more contemporary and precise view of the dose-response relationship, by including in the meta-analysis only RCTs published in the last 25 years (after 2000). This approach ensures relevance to current therapeutic and dietary management while minimizing variability associated with outdated BP measurement techniques or obsolete devices. We also restricted our inclusion criteria to studies where the intervention specifically targeted potassium intake and quantified it through 24-hour urinary potassium excretion, which is currently considered the “gold standard” for assessing potassium intake, despite its limitations. This avoided underestimation of potassium intake by other, more subjective methods of measurement based on patient recall and self-report, such as the Food Frequency Questionnaire (FFQ) and 24-hour recalls^61^, or urinary spots currently identified as unrepresentative^62,63^. Although the focus on recent studies guarantees current relevance, it may have led to the exclusion of some potentially useful older studies. This choice was deemed necessary to balance the need for high-quality data with practical constraints associated with study selection.

Contrary to our expectations, no other subgroup analyses (by dietary intervention, duration of intervention, study design or blood pressure measurement method — ambulatory or clinical)^64^ could be performed, due to the limited number of studies selected for this meta-analysis. Furthermore, we identified a significant gap in the literature: to our knowledge, no meta-analyses meeting our criteria have evaluated the impact of change in potassium intake on BP in patients with chronic kidney disease, a population at high cardiovascular risk. It is therefore essential to build RCTs measuring the impact of potassium intake (preferably by an increase in fruits and vegetables diet rather than potassium tablet supplementation) on BP in chronic kidney disease patients, in order to better understand its effects. The lack of correlation between kaliemia and kaliuresis^65–67^ and the expected benefit on BP and cardiovascular risk reduction supports the need for such trials in chronic kidney disease patients.

The dose-response analysis makes it possible to consider variations in the effect of modifying potassium intake, helping to refine the understanding and the assessment of the dose-response relationship. However, the limited number of studies (n=10) is certainly the greatest challenge to interpreting the models. The dose-response meta-analysis has nevertheless provided valuable information on the effect of potassium intake on BP variation, which remains the ignored ion in the sodium-potassium-blood pressure relationship^31^. Recent meta-analyses have highlighted the benefits of salt substitutes—formulated with reduced sodium chloride (NaCl) and enriched potassium chloride (KCl)—in lowering BP and reducing cardiovascular risk^17,18^. Unfortunately, these studies could not be included in our meta-analysis due to our strict inclusion criteria, which required a dietary intervention based solely on potassium modification. A simultaneous dietary intervention on sodium and potassium cannot dissociate the beneficial effect of increasing potassium intake and reducing sodium intake on BP. More rigorous regulation of both sodium and potassium intake could improve BP regulation thereby reducing cardiovascular morbidity and mortality. This dual regulation aligns with two ultimate objectives: improving both the quality and quantity of life for patients, and reducing healthcare costs associated with cardiovascular events and/or diseases, and the use of antihypertensive drugs^32^. This study could provide as a basis for the formulation of targeted nutritional recommendations and interventions adapted to specific populations, contributing to the prevention of cardiovascular disease and the management of hypertension. These objectives could only be achieved if the difficulties of implementing and enforcing long-term interventions to increase potassium intake are resolved. These difficulties include industrial opacity regarding the composition of processed foods^31^ and logistical challenges associated with 24-hour urine collection, which remains the standard measurement method but is restrictive. Efforts must be made to make the food industry and individuals aware of the cardiovascular risks associated with nutrition.

## Data Availability

All data is available on reasonable request from the author.

## Abbreviations and Acronyms

AIC: Akaike Information Criterion
BMI: Body Mass Index
BP: Blood Pressure
HP: High potassium
LP: Low potassium
PRISMA: Preferred Reporting Items for Systematic Reviews and Meta-Analyses
RCT: Randomized Controlled Trial
SBP: Systolic Blood Pressure
SE: Standard Error
SMD: Standardized Mean Difference

## Article information

## Author contributions

M Granal and A Gougeon formulated the review question and defined the inclusion and exclusion criteria. M Granal wrote the protocol and develop the search strategy in three specific databases. Data extraction was performed by M Granal and A Gougeon. M Granal analyzed data and drafted the paper. All authors read and approved the article. The corresponding author attests that all listed authors meet authorship criteria and that no other meeting the criteria have been omitted.

## Acknowledgment

We would like to thank Léonie Belot for her help in revising this article to improve its linguistic quality.

## Supplementary material

**Supplementary material S1:** PRISMA check list.

| Section and Topic | Item # | Checklist item | Location where item is reported |
| --- | --- | --- | --- |
| **TITLE** |  |  |  |
| Title | 1 | Identify the report as a systematic review. | Title |
| **ABSTRACT** |  |  |  |
| Abstract | 2 | See the PRISMA 2020 for Abstracts checklist. | Abstract |
| **INTRODUCTION** |  |  |  |
| Rationale | 3 | Describe the rationale for the review in the context of existing knowledge. | Introduction |
| Objectives | 4 | Provide an explicit statement of the objective(s) or question(s) the review addresses. | Introduction |
| **METHODS** |  |  |  |
| Eligibility criteria | 5 | Specify the inclusion and exclusion criteria for the review and how studies were grouped for the syntheses. | Methods - Search strategy and study selection |
| Information sources | 6 | Specify all databases, registers, websites, organisations, reference lists and other sources searched or consulted to identify studies. Specify the date when each source was last searched or consulted. | Methods - Search strategy and study selection |
| Search strategy | 7 | Present the full search strategies for all databases, registers and websites, including any filters and limits used. | Methods - Search strategy and study selection / supplementary material S2 |
| Selection process | 8 | Specify the methods used to decide whether a study met the inclusion criteria of the review, including how many reviewers screened each record and each report retrieved, whether they worked independently, and if applicable, details of automation tools used in the process. | Methods - Search strategy and study selection |
| Data collection process | 9 | Specify the methods used to collect data from reports, including how many reviewers collected data from each report, whether they worked independently, any processes for obtaining or confirming data from study investigators, and if applicable, details of automation tools used in the process. | Methods - Data extraction |
| Data items | 10a | List and define all outcomes for which data were sought. Specify whether all results that were compatible with each outcome domain in each study were sought (e.g. for all measures, time points, analyses), and if not, the methods used to decide which results to collect. | Methods - Data extraction |
|  | 10b | List and define all other variables for which data were sought (e.g. participant and intervention characteristics, funding sources). Describe any assumptions made about any missing or unclear information. | Methods - Data extraction |
| Study risk of bias assessment | 11 | Specify the methods used to assess risk of bias in the included studies, including details of the tool(s) used, how many reviewers assessed each study and whether they worked independently, and if applicable, details of automation tools used in the process. | Methods - Risk of bias assessment |
| Effect measures | 12 | Specify for each outcome the effect measure(s) (e.g. risk ratio, mean difference) used in the synthesis or presentation of results. | Methods - Data extraction / Statistical Analysis |
| Synthesis methods | 13a | Describe the processes used to decide which studies were eligible for each synthesis (e.g. tabulating the study intervention characteristics and comparing against the planned groups for each synthesis (item #5)). | Methods - Search strategy and study selection |
|  | 13b | Describe any methods required to prepare the data for presentation or synthesis, such as handling of missing summary statistics, or data conversions. | Methods - Search strategy and study selection / Data extraction |
|  | 13c | Describe any methods used to tabulate or visually display results of individual studies and syntheses. | Methods - Search strategy and study selection |
|  | 13d | Describe any methods used to synthesize results and provide a rationale for the choice(s). If meta-analysis was performed, describe the model(s), method(s) to identify the presence and extent of statistical heterogeneity, and software package(s) used. | Methods - Data extraction |
|  | 13e | Describe any methods used to explore possible causes of heterogeneity among study results (e.g. subgroup analysis, meta-regression). | Methods - Statistical Analysis |
|  | 13f | Describe any sensitivity analyses conducted to assess robustness of the synthesized results. | Methods - Statistical Analysis |
| Reporting bias assessment | 14 | Describe any methods used to assess risk of bias due to missing results in a synthesis (arising from reporting biases). | Methods - Statistical Analysis |
| Certainty assessment | 15 | Describe any methods used to assess certainty (or confidence) in the body of evidence for an outcome. | Methods - Statistical Analysis |
| **RESULTS** |  |  |  |
| Study selection | 16a | Describe the results of the search and selection process, from the number of records identified in the search to the number of studies included in the review, ideally using a flow diagram. | Results - Study selection and characteristics - Flowchart |
|  | 16b | Cite studies that might appear to meet the inclusion criteria, but which were excluded, and explain why they were excluded. | Results - Study selection and characteristics - Flowchart |
| Study characteristics | 17 | Cite each included study and present its characteristics. | Results - Study selection and characteristics - Table 1. Characteristics of the 10 RCTs included |
| Risk of bias in studies | 18 | Present assessments of risk of bias for each included study. | Results - Risk of bias assessment / Supplementary data 6 |
| Results of individual studies | 19 | For all outcomes, present, for each study: (a) summary statistics for each group (where appropriate) and (b) an effect estimates and its precision (e.g. confidence/credible interval), ideally using structured tables or plots. | / |
| Results of syntheses | 20a | For each synthesis, briefly summarise the characteristics and risk of bias among contributing studies. | Results - Dose-response meta-analysis / Dose-response prediction |
|  | 20b | Present results of all statistical syntheses conducted. If meta-analysis was done, present for each the summary estimate and its precision (e.g. confidence/credible interval) and measures of statistical heterogeneity. If comparing groups, describe the direction of the effect. | Results - Dose-response meta-analysis / Dose-response prediction |
|  | 20c | Present results of all investigations of possible causes of heterogeneity among study results. | Results - Publication bias assessment |
|  | 20d | Present results of all sensitivity analyses conducted to assess the robustness of the synthesized results. | Results - Publication bias assessment |
| Reporting biases | 21 | Present assessments of risk of bias due to missing results (arising from reporting biases) for each synthesis assessed. | Results - Publication bias assessment |
| Certainty of evidence | 22 | Present assessments of certainty (or confidence) in the body of evidence for each outcome assessed. | Results - Publication bias assessment |
| **DISCUSSION** |  |  |  |
| Discussion | 23a | Provide a general interpretation of the results in the context of other evidence. | Discussion |
|  | 23b | Discuss any limitations of the evidence included in the review. | Discussion |
|  | 23c | Discuss any limitations of the review processes used. | Discussion |
|  | 23d | Discuss implications of the results for practice, policy, and future research. | Discussion |
| **OTHER INFORMATION** |  |  |  |
| Registration and protocol | 24a | Provide registration information for the review, including register name and registration number, or state that the review was not registered. | Methods - Protocol and registration |
|  | 24b | Indicate where the review protocol can be accessed, or state that a protocol was not prepared. | Methods - Protocol and registration |
|  | 24c | Describe and explain any amendments to information provided at registration or in the protocol. | Methods - Protocol and registration |
| Support | 25 | Describe sources of financial or non-financial support for the review, and the role of the funders or sponsors in the review. | Article information - Sources of Funding |
| Competing interests | 26 | Declare any competing interests of review authors. | Article information - Disclosures |
| Availability of data, code and other materials | 27 | Report which of the following are publicly available and where they can be found: template data collection forms; data extracted from included studies; data used for all analyses; analytic code; any other materials used in the review. | Supplementary material |

**Supplementary material S2.** Search strategy for PubMed, Embase and Cochrane database. The global search equation developed made it possible to select all studies relating to the impact of modifying sodium and/or potassium intake on blood pressure which were the subject of two separate meta-analyses (CF PROSPERO protocol CRD42023440909).

| Database | Search items |
| --- | --- |
| PubMed |  |
| No. |  |
| #1 | "sodium"[MeSH Terms] OR "sodium"[tiab] OR "sodiums"[tiab] OR "Sodium, Dietary"[Mesh] OR "sodium chloride"[MeSH] OR "Sodium Chloride, Dietary"[Mesh] OR "NaCl"[tiab] OR "dietary sodium"[tiab] OR "sodium dietary"[tiab] OR "sodium chloride"[tiab] OR "salt"[tiab] |
| #2 | "potassium, dietary"[MeSH] OR "potassium" [MeSH] OR "potassium dietary"[tiab] OR "potassium" [tiab] OR "dietary potassium"[tiab] |
| #3 | "sodium to potassium ratio"[tiab] OR "Na/K ratio"[tiab] |
| #4 | "sodium glucose"[tiab] |
| #5 | "sodium bicarbonate"[tiab] |
| #6 | #1 OR #2 OR #3 NOT #4 NOT #5 |
| #7 | "blood pressure"[MeSH Terms] OR "blood pressure"[tiab] OR "blood pressure determination"[MeSH Terms] OR "blood pressure determination"[tiab] OR "arterial pressure"[MeSH Terms] OR "arterial pressure"[tiab] |
| #8 | "mortality"[MeSH Terms] OR "mortality"[tiab] OR "mortalities"[tiab] OR "lethal"[tiab] OR "lethalities"[tiab] OR "lethality"[tiab] OR "lethals"[tiab] OR "death"[MeSH Terms] OR "death"[tiab] OR "deaths"[tiab] OR "fatal"[tiab] |
| #9 | #7 OR #8 |
| #10 | "meta analysis"[Publication Type] OR "meta analysis as topic"[MeSH Terms] OR "meta analysis"[tiab] OR "meta-analysis"[tiab] OR "systematic review"[Publication Type] OR "systematic reviews as topic"[MeSH Terms] OR "systematic review*"[tiab] |
| #11 | "humans"[MeSH Terms] |
| **#12** | "renal insufficiency, chronic"[MeSH Terms] OR "chronic renal insufficiency"[tiab] OR "chronic kidney disease"[tiab] OR "renal insufficiency"[MeSH Terms] OR "renal insufficiency"[tiab] OR "kidney failure"[tiab] |
| **#13** | #11 OR #12 |
| **#14** | #6 AND #9 AND #10 AND #13 |
| **Result** | **841** |
| **Embase** |  |
| **#1** | "sodium"/exp OR "sodium chloride"/exp OR "sodium intake"/exp OR "sodium dietary":ab,ti OR "sodium chloride":ab,ti OR "sodium":ab,ti OR "NaCl":ab,ti OR "salt":ab,ti OR "dietary sodium":ab,ti OR "sodiums":ab,ti OR "salt intake"/exp |
| **#2** | "potassium dietary":ab,ti OR "potassium":ab,ti OR "dietary potassium":ab,ti OR 'potassium'/exp OR 'potassium intake'/exp |
| **#3** | "sodium to potassium ratio"/exp OR "sodium to potassium ratio»:ab,ti |
| **#4** | "sodium glucose":ab,ti |
| **#5** | "sodium bicarbonate":ab,ti |
| **#6** | #1 OR #2 OR #3 NOT #4 NOT #5 |
| **#7** | "blood pressure"/exp OR "blood pressure":ab,ti OR "blood pressure measurement"/exp OR "blood pressure determination":ab,ti OR "arterial pressure"/exp OR "arterial pressure":ab,ti |
| **#8** | "mortality"/exp OR "mortality rate"/exp OR "death"/exp OR "mortality":ab,ti OR "mortalities":ab,ti OR "lethal":ab,ti OR "lethalities":ab,ti OR "lethality":ab,ti OR "lethals":ab,ti OR "death":ab,ti OR "deaths":ab,ti OR "fatal":ab,ti |
| **#9** | #7 OR #8 |
| **#10** | "meta analysis"/exp OR "meta analysis (topic)/exp OR "meta analysis":ab,ti OR "meta-analysis":ab,ti OR "systematic review"/exp OR "systematic review (topic)/exp OR "systematic review":ab,ti |
| **#11** | 'human'/exp |
| **#12** | 'chronic kidney failure'/exp OR "chronic renal insufficiency":ab,ti OR "chronic kidney disease":ab,ti OR 'kidney failure'/exp OR "renal insufficiency":ab,ti OR "kidney failure":ab,ti |
| **#13** | #11 OR #12 |
| **#14** | #6 AND #9 AND #10 AND #13 |
| **Result** | **918** with date filtered to 2000 |
| **Cochrane** |  |
| **#1** | MeSH descriptor: [Sodium] explode all trees |
| **#2** | MeSH descriptor: [Sodium, Dietary] explode all trees |
| **#3** | MeSH descriptor: [Sodium Chloride] explode all trees |
| **#4** | MeSH descriptor: [Sodium Chloride, Dietary] explode all trees |
| **#5** | ("sodium dietary" OR "sodium chloride" OR "sodium" OR "NaCl" OR "salt" OR "dietary sodium" OR "sodiums"):ti,ab,kw |
| **#6** | MeSH descriptor: [Potassium] explode all trees |
| **#7** | MeSH descriptor: [Potassium, Dietary] explode all trees |
| **#8** | ("potassium dietary" OR "potassium" OR "dietary potassium"):ti,ab,kw |
| **#9** | ("sodium to potassium ratio", "Na/K ratio"):ti,ab,kw |
| **#10** | #1 OR #2 OR #3 OR #4 OR #5 OR #6 OR #7 OR #8 OR #9 |
| **#11** | ("sodium glucose"):ti,ab,kw |
| **#12** | ("sodium bicarbonate"):ti,ab,kw |
| **#13** | #10 NOT #11 NOT #12 |
| **#14** | ("blood pressure" OR "blood pressure determination" OR "arterial pressure"):ti,ab,kw |
| **#15** | MeSH descriptor: [Blood Pressure] explode all trees |
| **#16** | MeSH descriptor: [Blood Pressure Determination] explode all trees |
| **#17** | MeSH descriptor: [Arterial Pressure] explode all trees |
| **#18** | #14 OR #15 OR #16 OR #17 |
| **#19** | (mortalit* OR lethal* OR death* OR "fatal"):ti,ab,kw |
| **#20** | MeSH descriptor: [Mortality] explode all trees |
| **#21** | MeSH descriptor: [Death] explode all trees |
| **#22** | #19 OR #20 OR #21 |
| **#23** | #18 OR #22 |
| **#24** | MeSH descriptor: [Meta-Analysis as Topic] explode all trees |
| **#25** | MeSH descriptor: [Meta-Analysis] explode all trees |
| **#26** | MeSH descriptor: [Systematic Review] explode all trees |
| **#27** | MeSH descriptor: [Systematic Reviews as Topic] explode all trees |
| **#28** | ("meta analysis" OR "meta-analysis" OR systematic review*):ti,ab,kw |
| **#29** | #24 OR #25 OR #26 OR #27 OR #28 |
| **#30** | #13 AND #23 AND #29 |
| **Result** | **66** with date filtered to 2000 |

**Supplementary material S3.** Score Akaike of dose-response models of potassium intake on systolic blood pressure.

| Population | Linear | Quadratic | Cubic spline |
| --- | --- | --- | --- |
| All | 61,27 | 73,94 | 60,73 |
| Normotensive | 27,27 | 43,29 | 31,86 |
| Hypertensive | 37,29 | 51,44 | 42,14 |

**Supplementary material S4.**
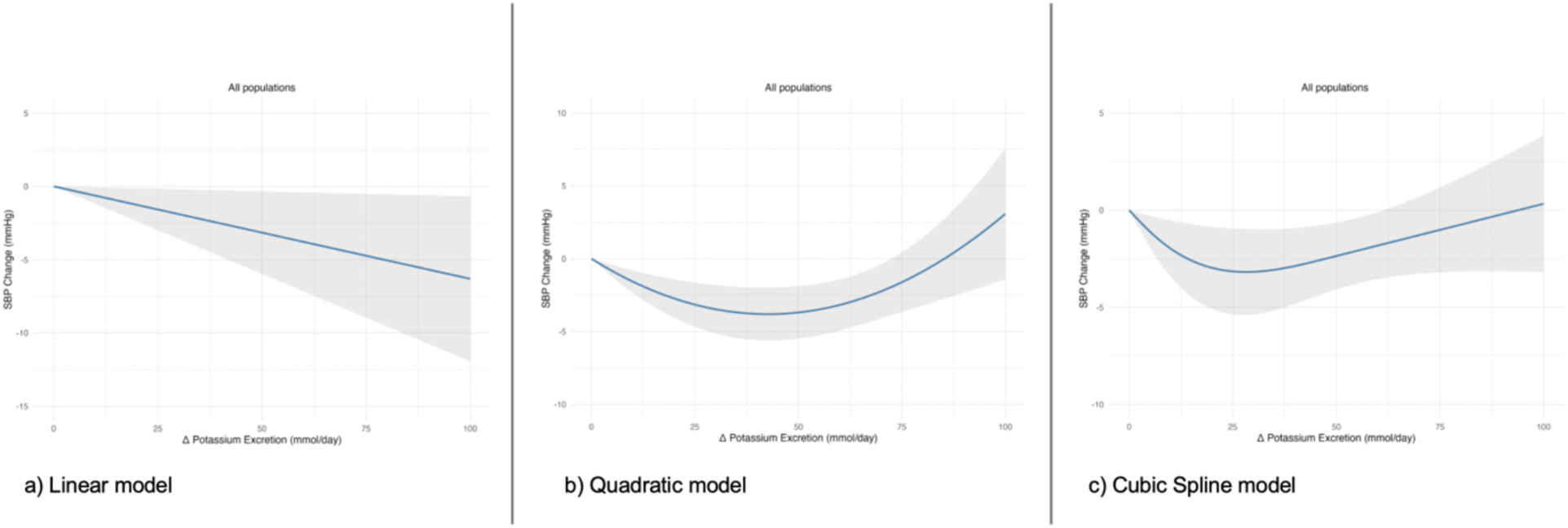
Dose-response meta-analysis of changes in systolic blood pressure (in mmHg) as a function of differences in urinary potassium excretion (in mmol/day) between the treatment group (potassium-supplemented group) and the control group with 95% confidence limits in both groups (normotensives and hypertensives; n=10 studies) with a) Linear model, b) Quadratic model, c) Cubic Spline model.

**Supplementary material S5.**
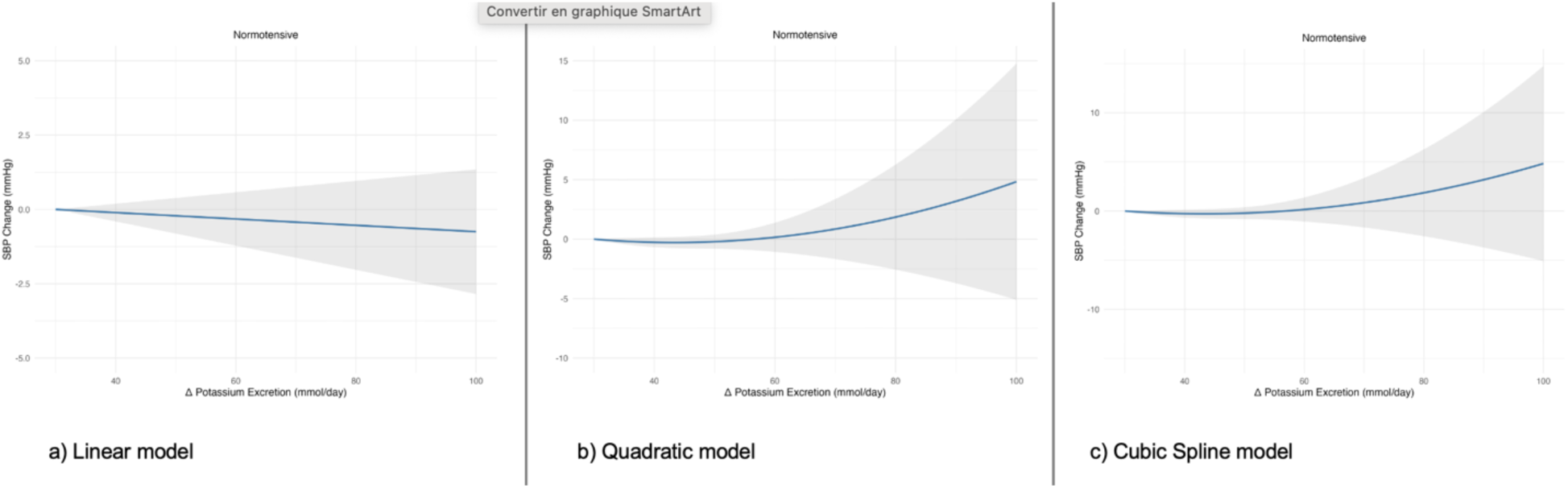
Dose-response meta-analysis of changes in systolic blood pressure (in mmHg) as a function of differences in urinary potassium excretion (in mmol/day) between the treatment group (potassium-supplemented group) and the control group with 95% confidence limits in the normotensive group (n=4) with a) Linear model, b) Quadratic model, c) Cubic Spline model.

**Supplementary material S6.**
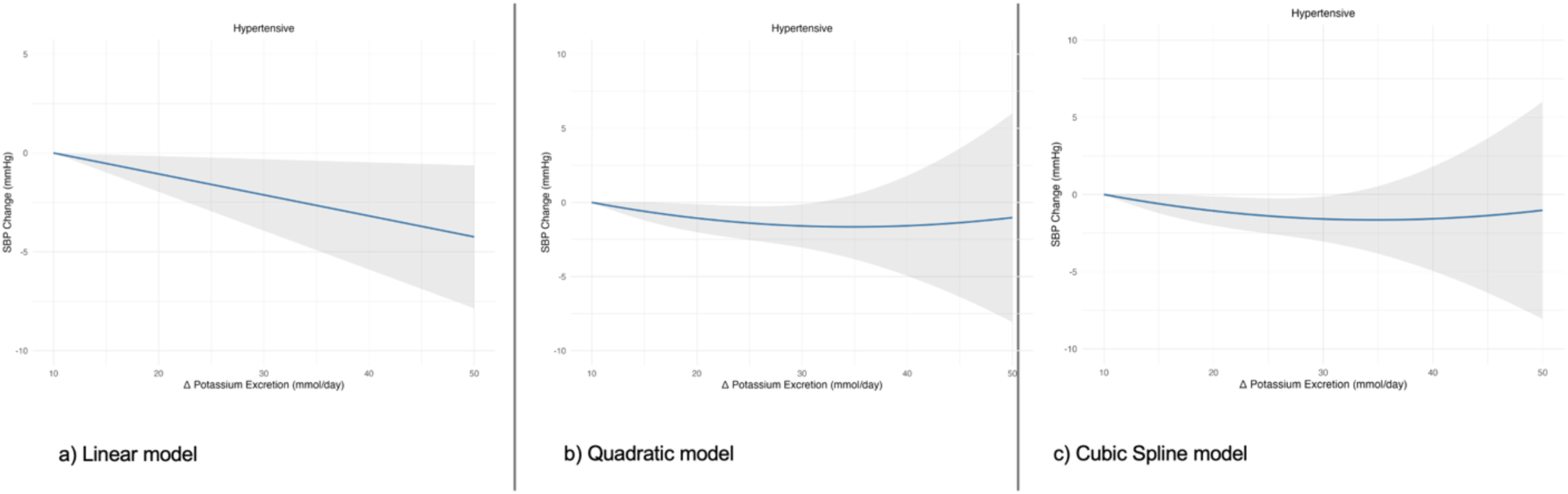
Dose-response meta-analysis of changes in systolic blood pressure (in mmHg) as a function of differences in urinary potassium excretion (in mmol/day) between the treatment group (potassium-supplemented group) and the control group with 95% confidence limits in the hypertensive group (n=4) with a) Linear model, b) Quadratic model, c) Cubic Spline model.

**Supplementary material S7.**
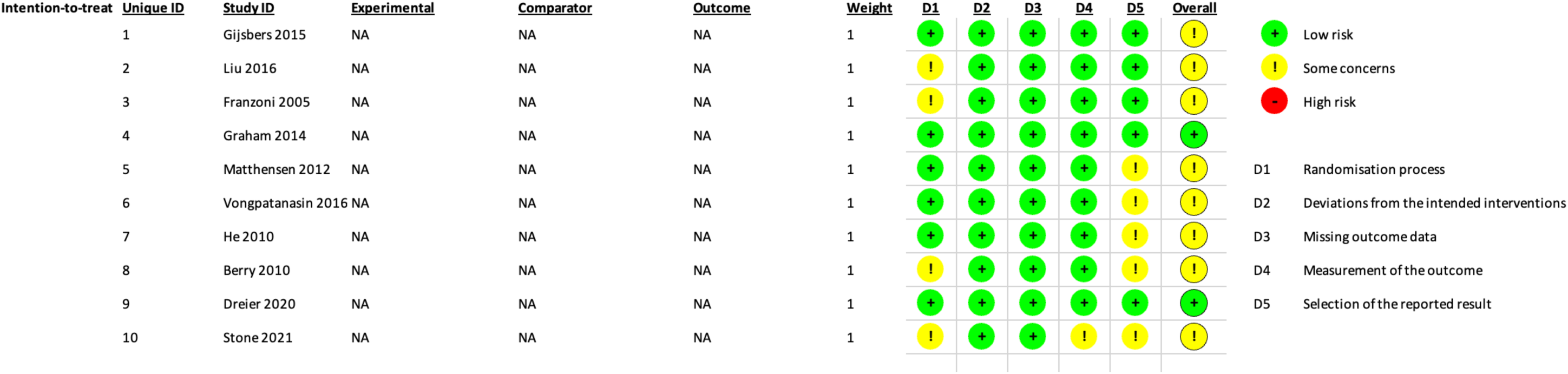
Details of risk of bias assessment.

